# The State of Household Handwashing by Mothers and Associated Risk Factors in Nepal: A Systematic Review

**DOI:** 10.1101/2022.06.06.22276036

**Authors:** Shalik Ram Dhital, Catherine Chojenta, Tanmay Bagade, Deborah Loxton

## Abstract

Approximately 53% of households do not use soap and water for handwashing. Mothers oversee their hygiene and are ideally placed to teach their children about handwashing with soap. This paper aims to determine the rates of household handwashing with soap by mothers in Nepal and explore the factors associated with the uptake of handwashing. A systematic literature search using PubMed/Medline, Embase, PsycINFO, CINAHL, and Grey literature was searched. Eligible articles were extracted and recorded then descriptive analysis was performed. The quality assessment of the article was done using STROBE guidelines. We screened 187 articles from database searching and 16 from additional grey literature. A total of 120 full-textt articles and records were retrieved to evaluate for inclusion in the review. We identified seven articles that met the criteria for inclusion. The included studies contained 32,010 participants from articles. Current handwashing rates by mothers were varied, ranging from 5% to 67%. The potential risk factors for a lack of handwashing were lack of knowledge, lack of handwashing facilities, and absence of regularities. The hygiene advocacy, provision of soap and water, family encouragement and support, the establishment of a handwashing strategy, and mothers’ participation in decision making are key possible solutions.

## Introduction

As the recent COVID-19 pandemic has highlighted, handwashing with soap is a practical, cheap, feasible, and straightforward way to prevent and control communicable diseases, especially in low-resource settings such as Nepal [1-3]. Handwashing with soap is a significant component of the prevention and control of skin infections, acute respiratory infections, and diarrhoea among children under five years [1,4,5]. This has been apparent at the global level with the campaigns related to COVID-19 transmission about prevention through handwashing with soap [6,7]. Reducing exposure to pathogens is a global health priority [9], demonstrated by the Sustainable Development Goals 6 (SDGs 2016–30) which have given priority to achieving universal access to all aspects of WASH by 2030 [10,11].

Handwashing at the household level is determined by several factors, such as knowledge of the importance of handwashing, risk communication, availability of water and soap, family ownership of soap, water and a fixed place for handwashing, installation of tippy taps (a hands-free way to wash hands – especially appropriate for water-scarce rural areas – which is operated by a foot lever and may increase the rate of handwashing with soap), perceived cost, and an individual’s ‘busy schedule and tiredness[12-17]. Household handwashing in Nepal is also influenced by context-specific handwashing policies, strategies and guidelines, as well as geographical and environmental factors [18-20]. Handwashing within households as a standard practice is still not widespread throughout the country. Despite these facilitators and challenges, handwashing remains a key method of reducing communicable disease rates [4,21].

In Nepal, mothers are primarily caregivers for their children. They teach children at home about handwashing with soap, and managing handwashing facilities with family members support [22,23]. Handwashing with soap promotion campaigns have a positive impact on children’s health [24]. Handwashing with soap practice provides children with safe and clean home environments [25]. Family members, such as the husband, father-in-law, and mother-in-law, can hand wash with soap by buying soap, and managing water and fixed places for handwashing. In this discourse, mothers are important persons, as they can be role models in the household.

The five key critical moments recommended to wash hands are: before eating or preparing food; before breastfeeding and feeding children; after defaecating or using the toilet; after cleaning a child faeces or handling nappies; and after touching a source of contamination.^1^ In Nepal, the overall handwashing knowledge of mothers was 60% in 2014 [26]. The rate of handwashing with soap by mothers before handling food was 67% after a three-month awareness program in Kavre district, while the baseline survey rate of handwashing with soap was only 5% in 2015 [27]. A study conducted in Rolpa district showed the self-reported prevalence of handwashing was 8% at baseline, 96% after a handwashing intervention, and 77% at follow-up, 30 months after the intervention [28]. However, the rates of handwashing with soap before preparing food, before feeding children or breastfeeding, and after cleaning children’s bottoms was between 6-22% in Nepal [26]. The application of existing knowledge regarding handwashing, especially before child feeding and breastfeeding and after the disposal of child faeces, is a challenge and barrier to good hygiene practices.

With such varying rates of handwashing with soap during critical moments, it is essential to examine this issue in Nepal. This paper aims to determine the household handwashing rates with soap by mothers in Nepal and explore the factors associated with the uptake of handwashing.

## Materials and Methods

### Search strategy and selection criteria

This systematic review adopted the Preferred Reporting Items for Systematic reviews and Meta-Analysis (PRISMA) checklist [29]. Published literature was searched in the following databases: PubMed/Medline, Embase, PsycINFO, and Cumulative Index to Nursing & Allied Health Literature (CINAHL). A manual search for articles was also conducted in Google Scholar. Keywords included *handwashing, situation, households, mothers, children, determinants, knowledge, soap, health education*, and *Nepal*. The *g*rey literature (for example government reports, project reports, working papers, technical reports, and unpublished theses) was searched using keywords that were the same as those used to search the peer-reviewed literature. Relevant papers were also hand-searched. Articles were included if (i) the study was conducted in Nepal, (ii) information was collected from mothers, and (iii) the study was published in English. No limits were placed on the dates of data collection or publication. Articles were excluded if they were animal studies, non-mother samples, protocol papers, systematic reviews, abstracts only, or editorials. The search was limited to November 2019. The details of the screening of articles are in the PRISMA flow chart (Figure 1).

**Figure 1.**
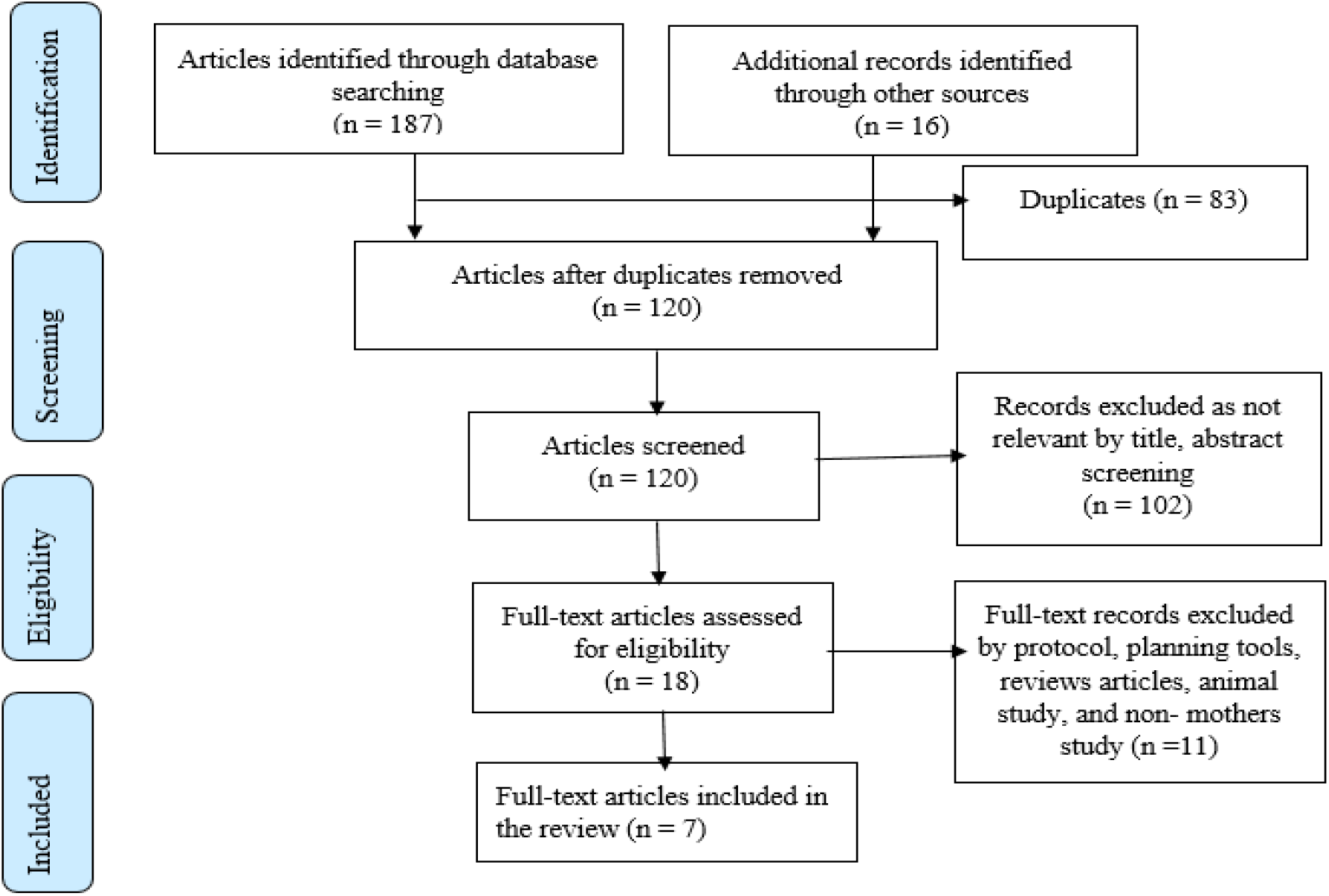
PRISMA flow diagram

### Data extraction, analysis and quality assessment

All eligible articles and records were extracted and recorded in an Excel spreadsheet. Extracted information included author, year of publication, study design, participants, age group, study place, study periods, and outcome measures.

Descriptive analysis was performed for this review paper. Two reviewers (SRD and TB) finalized the list of articles and records that would be included. Meta-analysis was impossible due to the low number of studies identified and the heterogeneity of the outcome measures. A quality assessment was done (appendix B) using the Strengthening the Reporting of Observational Studies in Epidemiology (STROBE) guidelines [30]. All required fields of the guidelines were completed by the first reviewer (SRD), and cross-verification was done by the second reviewer (TB). Once the extraction of eligible studies was completed, a narrative synthesis was made to provide evidence about handwashing with soap by mothers. The characteristics recorded for all eligible articles comprised of the first author’s name, publication year, study design, study population, sample size, study periods, and key findings. The statement of the main findings and strengths and limitations were carefully reviewed and reported in the conclusion section. The main results were presented to answer the study aim.

## Results

### Study flow and characteristics of included studies

Initially, 187 articles were identified from the database search, and 16 records through the grey literature search. Of the total 203 records, 83 were excluded because of duplication. The remaining 120 were screened, and further records (n=102) were excluded due to exclusion criteria of irrelevant titles or abstracts. A total of 18 records were assessed for eligibility. Then, 11 full text articles and records were excluded by protocol, planning tools, review articles, animal study and non-mothers’ studies. Finally, seven full- text articles met all inclusion criteria (Table 1). Of the seven studies found, three studies described randomized control trials (RCT), conducted in Kavre, Kathmandu, and Chitwan, Makwanpur, and Nuwakot districts. Three studies were cross sectional, conducted in a rural and urban settings. One study described a cohort study, conducted in Sarlahi district. A summary of the results of the keywords searches can be found in Appendix A.

**Table 1.** Summary of methods

| **First author, Year** | **Study design** | **Sample size** | **Study periods** | **Main findings** |
| --- | --- | --- | --- | --- |
| Kafle S,2018[31] | Cross-sectional | 178 | Not specified | About 79% of the mothers had good knowledge about WASH, considering access to an improved water source, sanitary toilets and handwashing with soap indicators. |
| Kandel P,2017 [34] | Cross-sectional | 1,421 | Not specified | Availability of soap, water, and a fixed place for handwashing was significantly associated with lower rates of faecal contamination in water sources. |
| Gautam OP,2017 [27] | RCT | 239 | 3 months | Handwashing at home before feeding the child/children and before eating increased from 5% to 67%. |
| Miller LC, 2017[35] | RCT | 1,011 | 48 months | The frequency of usage of soap and availability of water at home was higher among mothers than fathers. The participatory Community |
|  |  |  |  | Development program was an effective way of increasing hygiene in households. |
| Langford R,2013[32] | RCT | 88 | 6 months | Approximately 21% of mothers washed their hands with soap after defaecation and 14% after cleaning baby's bottom. No data were obtained about handwashing during/before cooking or feeding the child in the non-intervention group. Intervention groups (baseline and post-intervention) showed that mothers washed their hands after defaecation (96% and 100%), after cleaning baby's bottom (82% and 100%), before cooking (12% and 71%), before feeding children (26% and 62%) and before eating (14% and 60%), respectively. |
| Rhee V, 2008[24] | Cohort study | 23,662 | 40 months | Prevalence of maternal handwashing with soap before handling their infants was 15%. Maternal handwashing with soap was associated with significantly lower rates of neonatal mortality. |
| Osrin D, 2002[33] | Cross-sectional | 5,411 | Not specified | Approximately 50% of mothers who attended a birth had washed their hands with soap. |

### Rates of household handwashing with soap by mothers in Nepal

Maternal rates of handwashing with soap varied during different critical moments in the eligible studies. A cross-sectional study conducted by Kafle and Pradhan in Makwanpur district among 178 mothers in 2018 reported that approximately 43% of mothers washed their hands with soap at critical moment while 79% of mothers had good knowledge about WASH [31]. Gautam et al conducted a RCT between October 2012 and December 2013 using structured observations of handwashing with soap among 239 mothers with children aged 6–59 months in Kavre showed that handwashing with soap before eating and feeding a child was 67% after a food hygiene campaign, which was significantly higher than the with 5% who undertook handwashing with soap before eating or feeding a child at baseline [27]. Langford and Panter-Brick conducted an RCT in the slum area of Kathmandu in 2013. They reported that the handwashing with soap rates were 100% after using the toilet and after cleaning children’s bottoms, 71% before cooking food, 62% before child feeding, and 60% before eating in the handwashing intervention arms of the study; while the results for the control arms were 91% after using the toilet, 84% after cleaning children’s bottoms, 19% before child feeding, 2.3% before cooking and zero percent before eating [32]. An observational prospective cohort study carried out in 2008 in Sarlahi district showed that mothers’ handwashing with soap prior to handling infants was only 15% [24] Likewise, another retrospective cross-sectional study conducted in the Makwanpur district among 5,411 mothers aged 15–49 years who had live births in the previous year in 2002 showed that approximately 50% of the birth attendants washed their hands prior to attending the deliveries [33].

These results indicated that handwashing with soap rates had a wide variation depending on the areas, circumstances, education, and critical time point being assessed, with the majority of results showing far less than optimal rates of handwashing with soap across the country.

### Factors associated with maternal handwashing in Nepal

A cross-sectional study carried out among randomly selected mothers from 178 households in Makwanpur district in 2018 found maternal knowledge and the household wealth index affect handwashing with soap practices [31]. In 2017, Kandel et al conducted a cross-sectional study using the 2014 Multiple Indicator Cluster Survey (MICS) dataset among 1,421 households’ mothers and reported that the faecal contamination of water was associated with the availability of adequate handwashing facilities with soap and water [34]. As mentioned above, an RCT carried out in a rural village in Kavre district in 2015 found mothers’ participation in Health Mothers’ Group meetings, and motivation through family support and rewards are enabling factors for handwashing with soap, while poor participation, demotivation, and punishment decrease handwashing with soap [27]. This study further found that an integrated health promotion campaign increased the rate of handwashing with soap. Miller et al conducted a RCT in Chitwan, Nawalparasi, and Nuwakot districts reported that women who attained higher-level of education had more frequent use of soap during handwashing compared with women with no education in 2017, and the participatory community development program was an effective way of increasing hygiene practices [35].

A study carried out in Kathmandu in 2013 showed that family and community beliefs, such as believing handwashing with soap is unnecessary, being unsure about good health after using soap, and the financial burden of buying soap, are barriers to effective handwashing with soap [32]. This study found that the community strongly believed that keeping children clean all the time causes illness, and some infectious diseases are caused by cold weather, fever, and evil spirits, not due to poor handwashing practices. A strong misconception is that handwashing with soap may not be necessary to prevent diseases that can have adverse health effects on people, especially the vulnerable populations such as children, pregnant mothers, and the elderly. A further barrier to handwashing with soap is that household members need to spend money on food, and which takes priority over soap.

A cohort study carried out in 2008 in the plains region in Sarlahi district of Nepal indicated that the possible factors associated with maternal handwashing were lack of education, absence of a toilet at home, and having low-birthweight babies [24]. A study conducted by Osrin and colleagues in the Makwanpur district among 4,511 mothers in 2002 found lack of knowledge about the importance of handwashing and hygiene, especially during breastfeeding and birth attendance, is a possible factor affecting effective handwashing in Nepal [33]. This review’s key summary results are shown in Table 1, including the first author’s name, publication year, study design, sample size, study periods, and main findings.

## Discussion

This systematic review aimed to determine the household handwashing rates with soap by mothers in Nepal and explore the factors associated with the uptake of handwashing.

Nepalese mothers typically take the primary caring role for children and family members and clean the home. These roles are traditionally established in Nepal. Handwashing should perform handwashing; however, mothers feel more pressure to provide safe hygiene for their children [36]. The mother’s role of caring for children is socially constructed in Nepal and is not counted as work in the home [37].

The first and foremost issue for general family education on handwashing is improving maternal handwashing knowledge. The NDHS 2016 results showed that handwashing facilities with soap and water at the household level were 47%, whereas the MICS 2014 found that mothers’ handwashing with soap knowledge varied depending on the specific critical moment being observed. For example, after cleaning children’s bottoms or changing nappies, handwashing rates of 6% were found compared to before meals, when 92% washed their hands with soap [26,38].

This review determined that maternal knowledge about the importance of handwashing with soap before eating or child feeding was higher after food hygiene intervention [27]. compared to after cleaning a child who has defaecated [32], and this is one of Nepal’s significant public health challenges [26]. Similarly, less than half of the mothers washed their hands with soap who attended childbirth[33]. The reason behind such low rates of handwashing by mothers may be related to the level of health knowledge about the threat and severity of not washing hands, unavailability of soap and water, financial crisis, and the cultural belief that communicable diseases (for example diarrhea) exist because of colds, fever, or evil spirits, rather than lack of handwashing [32]. In Nepal, mothers who are poor and those in rural and hard-to-reach areas remain most vulnerable to communicable diseases, due to inadequate access to health education and handwashing services [26]. The gap between handwashing knowledge and access to handwashing facilities with soap and water is a further challenge in Nepal [31].

This review highlighted that the significant factors and barriers to handwashing with soap are lack of knowledge, contrary beliefs, unavailability of soap and water, lack of money, demotivation of mothers, and low mother’s participation in planning and household decision-making. Maternal knowledge about handwashing is associated with handwashing practices [39]. Availability of soap, clean water, fixed places, adequate time, and family, as well as community support, encourage mothers to wash their hands while the absence of such factors are barriers to uptake of handwashing [16,40]. These may be overcome by family support such as the construction of fixed places for handwashing near or inside the home, buying soap, and provision of convenient improved water sources. Timely and accurate health education for household members by health workers contributes to counteracting barriers. For example, if mothers know about the need for handwashing at critical moments, they are likely to put this knowledge into practice. This statement is supported by a previous study carried out in Korea in 2013, which argued that providing health education improved handwashing [41]. Mothers are the key role models who can change household handwashing conditions. The results of this review indicate that factors affecting handwashing were similar to a 2015 Nepal-specific study of four various plains districts (Mahottari, Siraha, Saptari and Sarlahi), which showed that participating mothers were more likely to wash their hands with soap when their hands looked dirty, to have their hands soft and smelling good, and to keep their dignity [42]. This review is further supported by a previous case study report in Nepal which found that the significant challenges to proper handwashing with soap at the household level by mothers were providing high-quality education to mothers to increase health literacy about risks and threats, and to establish habits of utilizing available resources, such as soap, water, and handwashing stations. Further potential factors which determine handwashing with soap by mothers at the household level are family roles and responsibilities, household structure, geography, and climate [28].

A critical factor in improving the rate of handwashing with soap is changing the mindset of individuals in the community. However, the changing attitudes and behaviours is complex, and can be hampered by several factors. For example, slow economic growth and societal inequalities are challenges for the development of a health system, health promotion, and the practice of handwashing. It became clear during this review that high-quality and results-oriented public health and health promotion campaigns are required to overcome such factors at the household level regarding handwashing by mothers and in turn the whole community. Context-specific handwashing promotional materials or facilities for mothers as well as families and households, timely high-quality health education sessions on handwashing, and policy, guidelines and strategies documents are needed. Simple, affordable, feasible and practical interventions are required at the local level. During planning and designing handwashing messages and interventions, the local cultural, social, economic, geographical, and other contextual factors should be considered [43]. Community-based health promotion actions are recommended to deliver the handwashing message through processes of advocacy, services, and policy formulation and application approaches [44]. This review further recommends conducting multivariate and multi-level analyses on handwashing with soap and water, and Nepal’s sanitation facilities. The quality and scope of the studies in this review suggest that participant observation studies on handwashing with soap should be undertaken. Priorities for future research should consider the effects of health education interventions, human resources, and high-quality handwashing facilities.

The limitations of this review must be acknowledged. Firstly, not all household-level handwashing rates and facilities were covered. Secondly, none of the included studies reported the availability of soap and water at the household level for mothers in Nepal. Thirdly, information about corrective measures for effective improvement of handwashing knowledge and behavioural change through community efforts was lacking.

## Conclusions

The previous studies suggested that handwashing is a cost-effective and affordable health promotion initiative to reduce infectious diseases like skin infections, acute respiratory infection and diarrhoea and most recently COVID-19 [4,6,21,45], thus supporting the claim of the importance of handwashing with soap. Even though Nepal has adopted SDGs 6 (2016-2030), still the progress of improving handwashing with soap is stable. The handwashing with soap rates by family members varied by place, person, and moment in Nepal. Further studies about the washing with soap before breastfeeding, after child faeces cleaning, and before preparing meals, are required. Low handwashing rates with soap were due to lack of knowledge, poverty, culture, and lack of soap and water at home. Addressing hygiene at the household level requires a thorough understanding of the factors related to mothers’ and family members’ knowledge and attitudes, sociodemographic, cultural, and economic factors. Water and soap are the pre-requisites for good hygiene and their availability is necessary; but much more needs to be done if household handwashing is to become a habit among mothers, children and other family members.

Water and soap are the pre-requisites for good hygiene, and their availability is necessary. However, much more needs to be done if household handwashing is to become a habit among mothers, family members, and children. A few policies related to WASH in Nepal have focused on health and hygiene, but there are no handwashing policies or strategies. Ensuring handwashing with soap is a regular habit remains a challenge in Nepal.

Studies related to handwashing with soap included in this review showed that an adequate supply of soap and water and construction of fixed place for handwashing including proper health education can help mothers to practice handwashing. The family income, husband and mother-in-law’s support for water fetching, construction of water sources near homes, construction of handwashing stations, and buying soap helps mothers to introduce and continue regular appropriate handwashing. Varies rates of handwashing with soap by mothers during critical moment remains a challenge and the participatory community development and health promotion programs require to improve knowledge about the risks and the provision of soap and water at home. Therefore, it is essential to conduct further studies on the availability of improved water sources, sanitary toilets, and handwashing with soap facilities at the household level. The roles of non-health stakeholders for improving handwashing with soap practices, and factors associated with the barriers and challenges to handwashing with soap in Nepal, also require study.

## Data Availability

After the search dataset, the information was obtained and kept in the supporting list of the Manuscript.

## Abbreviations

MICS: Multiple Indicator Cluster Survey;
PRISMA: Preferred Reporting Items for Systematic reviews and Meta-Analysis;
RCT: Randomised Control Trails;
STROBE: Strengthening the Reporting of Observational Studies in Epidemiology;
SDGs: Sustainable Development Goals;
WASH: Water, Sanitation and Hygiene.

## Acknowledgments

We would like to thank Dr. Ryan O’Neill from the University of Newcastle’s Centre for Women’s Health for providing an editorial review.

## Ethics approval and consent to participate

Not applicable

## Consent for publication

Not applicable

## Author’s contribution

Dr Shalik Ram Dhital conceptualized, coded data, extracted findings of articles and prepared the draft manuscript. Dr Tanmay Bagade gave input for making this manuscript worthy. Professor Deborah Loxton and Dr Catherine Chojenta contributed through academic and scientific feedback and suggestions with necessary modifications. All authors read and approved the final manuscript.

## Supplementary documents

## Appendix A. Summary of article search strategy

**Database(s): Ovid MEDLINE(R) and Epub Ahead of Print, In-Process & Other Non-Indexed Citations and Daily 1946 to November 07, 2019**

**Table I.**

| # | Searches | Results |
| --- | --- | --- |
| 1 | Handwashing.mp. or Hand Disinfection/ | 6303 |
| 2 | Situation.mp. | 149716 |
| 3 | households.mp. or Family Characteristics/ | 48061 |
| 4 | 1 or 2 or 3 | 202641 |
| 5 | Mothers/ or Mothers.mp. | 150026 |
| 6 | Children.mp. or Child/ | 2012115 |
| 7 | 5 or 6 | 2104630 |
| 8 | Determinants.mp. | 151783 |
| 9 | Knowledge/ or Health Knowledge, Attitudes, Practice/ | 116543 |
| 10 | Soap.mp. or Soaps/ | 5976 |
| 11 | Health education.mp. or Health Education/ | 85051 |
| 12 | 8 or 9 or 10 or 11 | 343876 |
| 13 | Nepal.mp. or Nepal/ | 10711 |
| 14 | 4 and 7 and 12 and 13 | 58 |

**Database(s): PsycINFO 1806 to October Week 4 2019 Search Strategy:**

**Table I.**
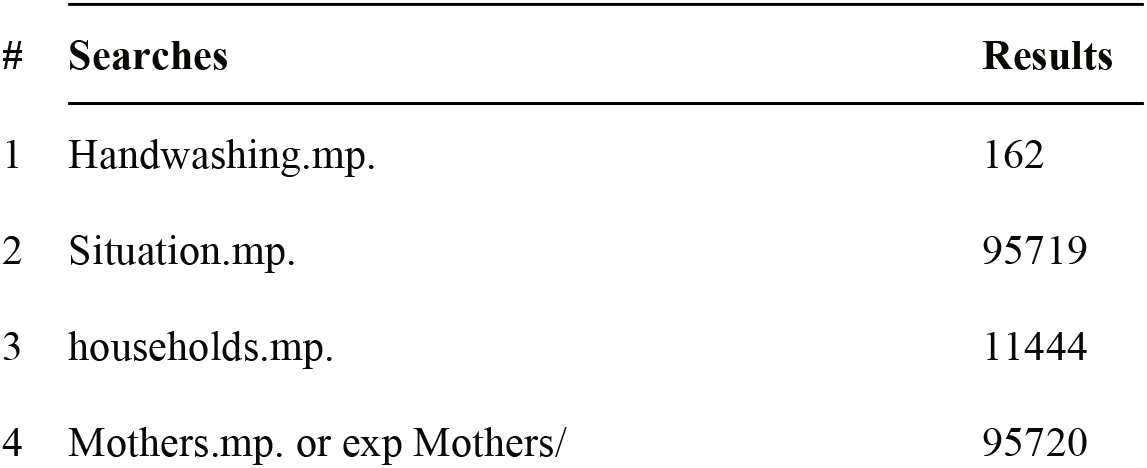

**Database(s): Embase 1947 to 9 Nov 2019 Search Strategy:**

**Table I.**
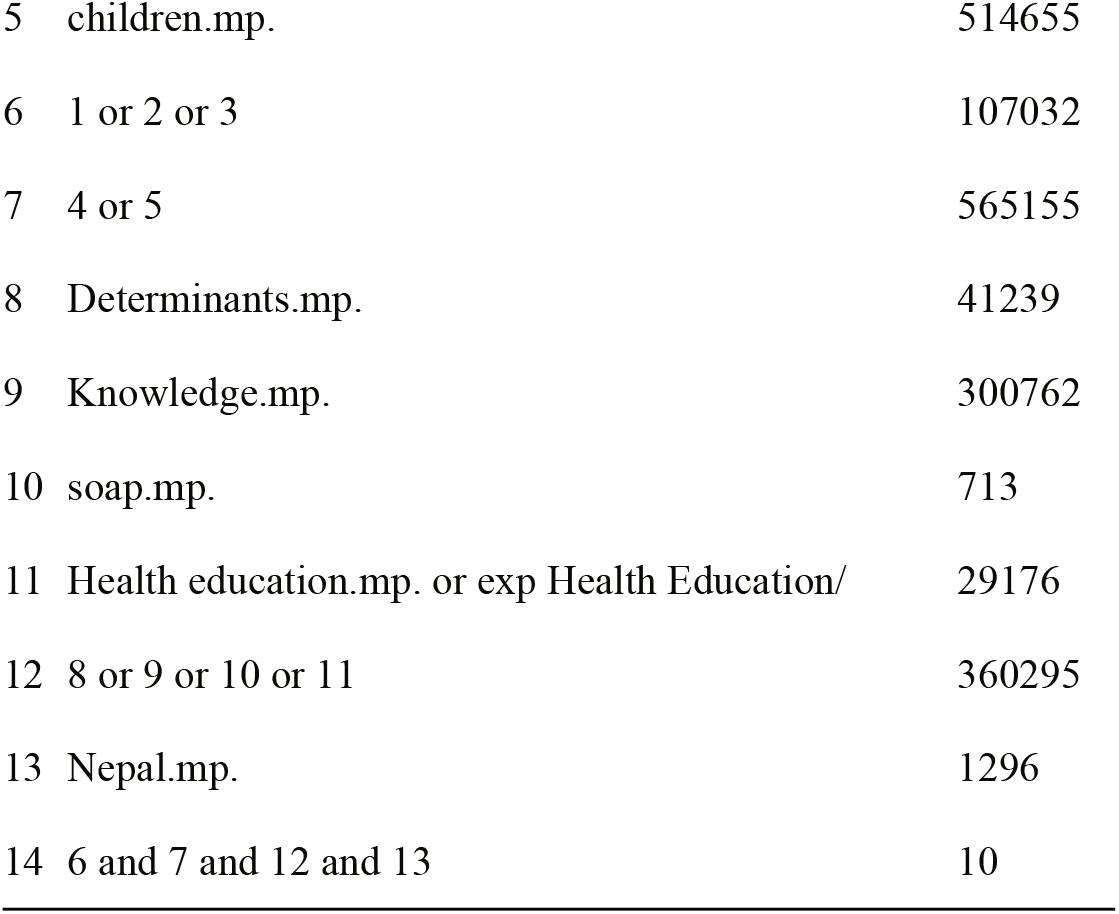

**Database(s): CINAHL until 9 Nov 2019**

**Table I.**

**Database(s): Embase 1947 to 9 Nov 2019 Search Strategy:**
| # | Searches | Results |
| --- | --- | --- |
| 1 | Handwashing.mp. or handwashing/ | 13721 |
| 2 | Situation.mp. | 246330 |
| 3 | households.mp. or household/ | 56559 |
| 4 | 1 or 2 or 3 | 314617 |
| 5 | Mothers.mp. or mother/ | 219548 |
| 6 | Children.mp. or child/ | 2341181 |
| 7 | 5 or 6 | 2476177 |
| 8 | Determinants.mp. or "social determinants of health"/ | 187215 |
| 9 | Knowledge.mp. | 873291 |
| 10 | Soap/ or Soap.mp. | 9349 |
| 11 | Health education.mp. or health education/ | 120991 |
| 12 | 8 or 9 or 10 or 11 | 1157688 |
| 13 | Nepal.mp. or Nepal/ | 13087 |
| 14 | 4 and 7 and 12 and 13 | 76 |

**Database(s): CINAHL until 9 Nov 2019**

**Table I.**
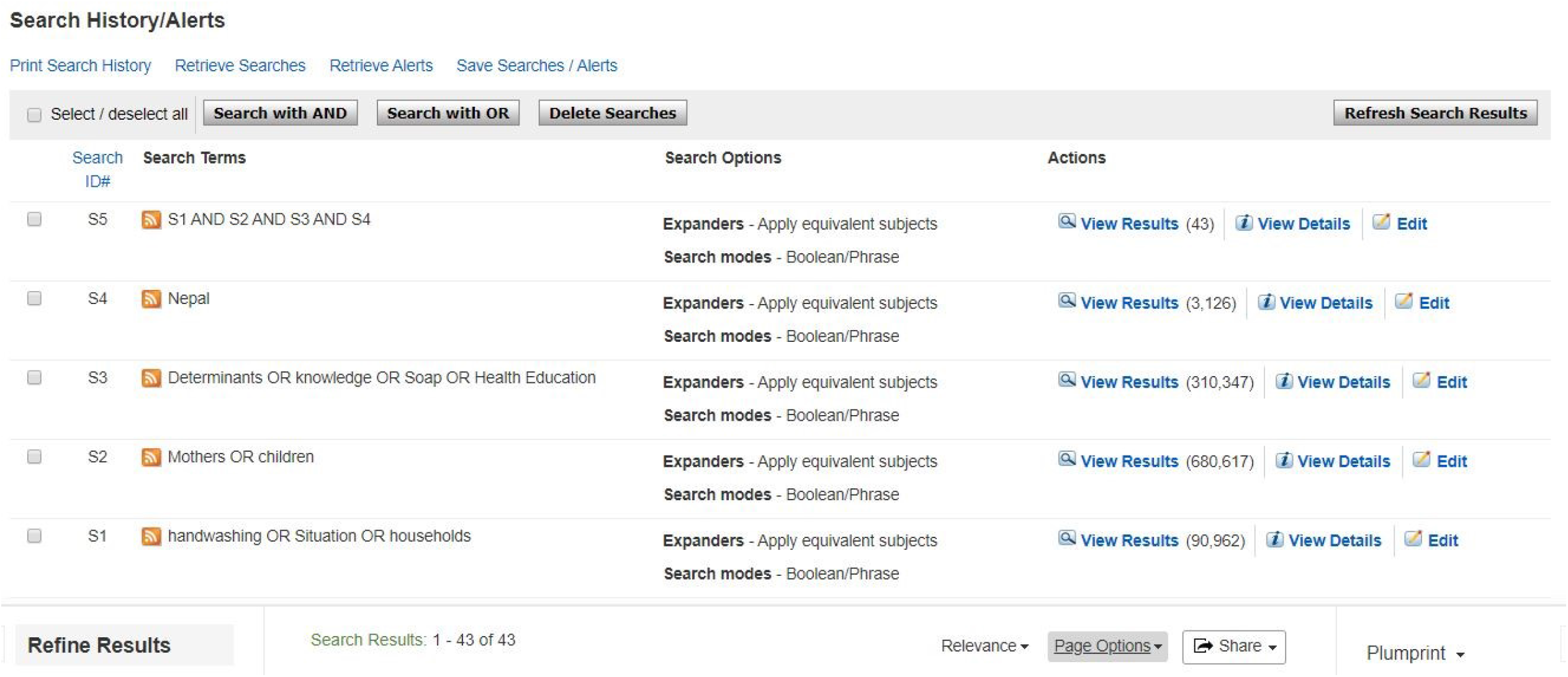

**Appendix B**. Strengthening the reporting of observational studies in epidemiology (STROBE) checklist, including paper grading

**Table I.**
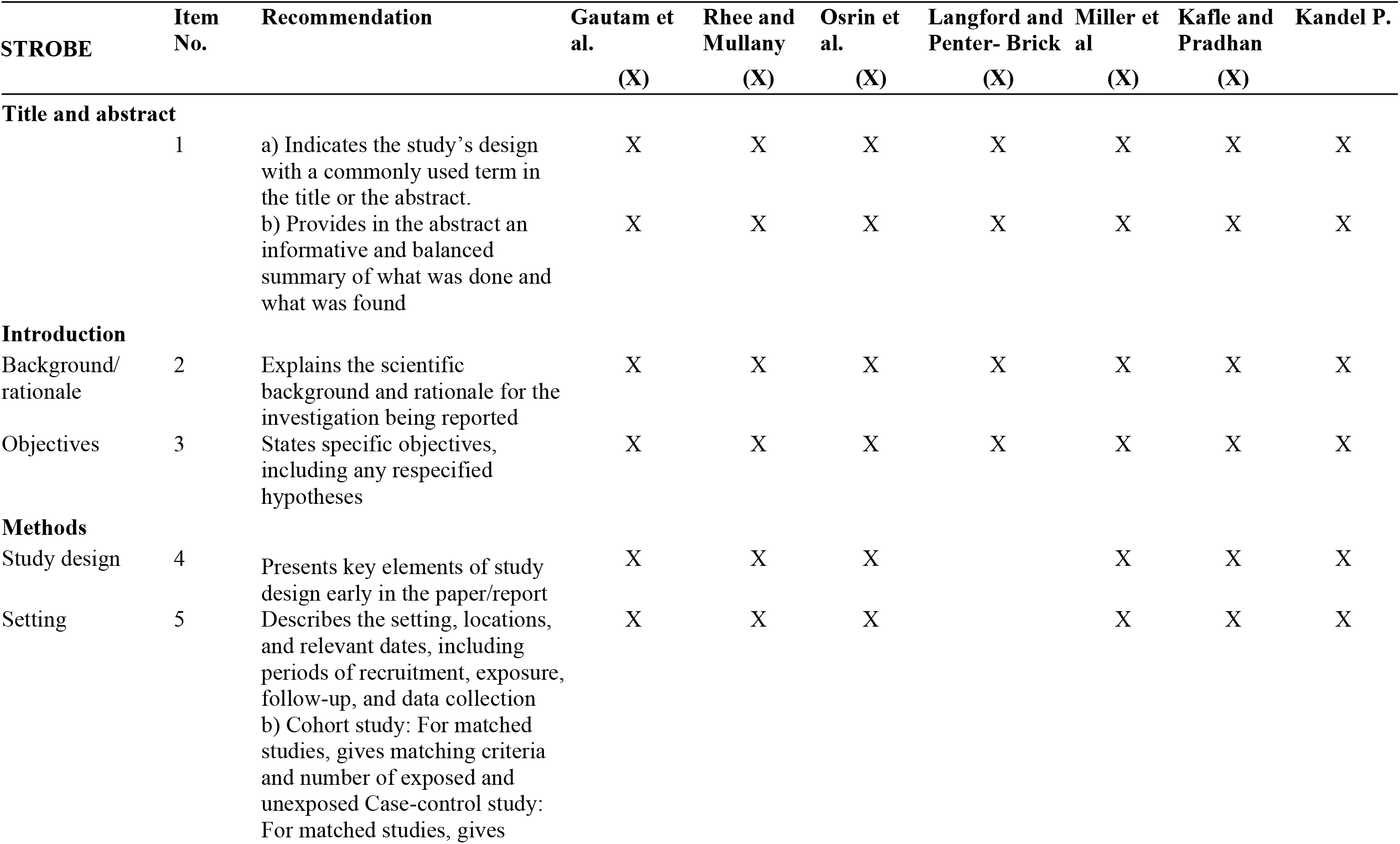

**Table I.**

|  |  | matching criteria and the number of controls per case |  |  |  |  |  |  |  |
| --- | --- | --- | --- | --- | --- | --- | --- | --- | --- |
| Variables |  | Clearly defines all outcomes, exposures, predictors, potential confounders, and effect modifiers. Gives diagnostic criteria, if applicable | X | X | X |  | X | X |  |
| Data sources/<br>measurement | 8 | For each variable of interest, gives sources of data and details of methods of assessment (measurement). Describes comparability of assessment methods if there is more than one group | X | X |  | X | X | X |  |
| Bias | 9 | Describes any efforts to address potential sources of bias | X |  | X |  | X |  |  |
| Study size | 10 | Explains how the study size was arrived at | X | X | X | X | X |  | X |
| Quantitative variables | 11 | Explains how quantitative variables were handled in the analyses. If applicable, describes which groupings were chosen and why | X | X | X | X | X |  |  |
| Statistical methods | 12 | a) Describes all statistical methods, including those used to control for confounding<br>b) Describes any methods used to examine subgroups and interactions<br>c) Explains how missing data were addressed | X | X |  | X | X | X |  |

**Table I.**
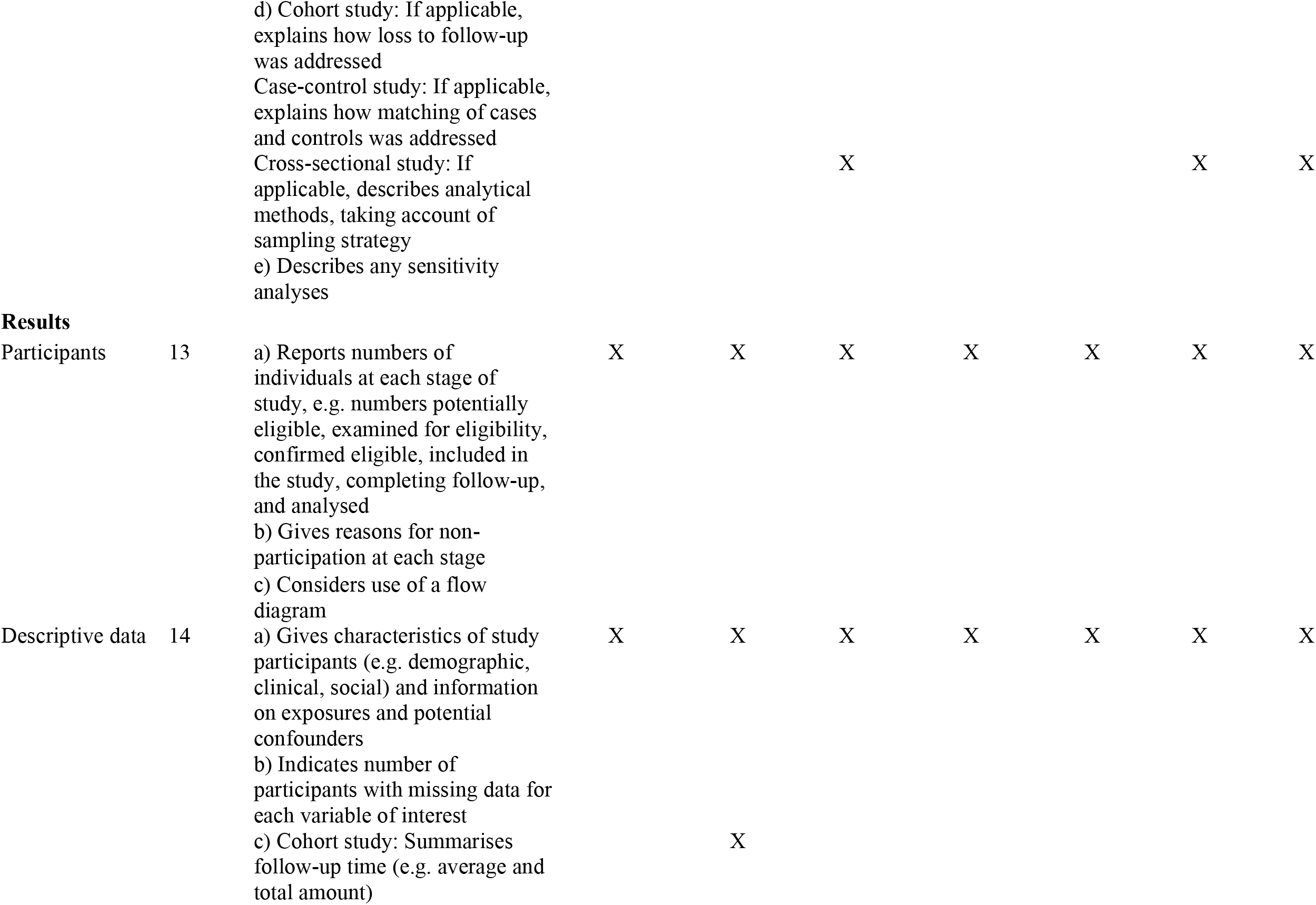

**Table I.**
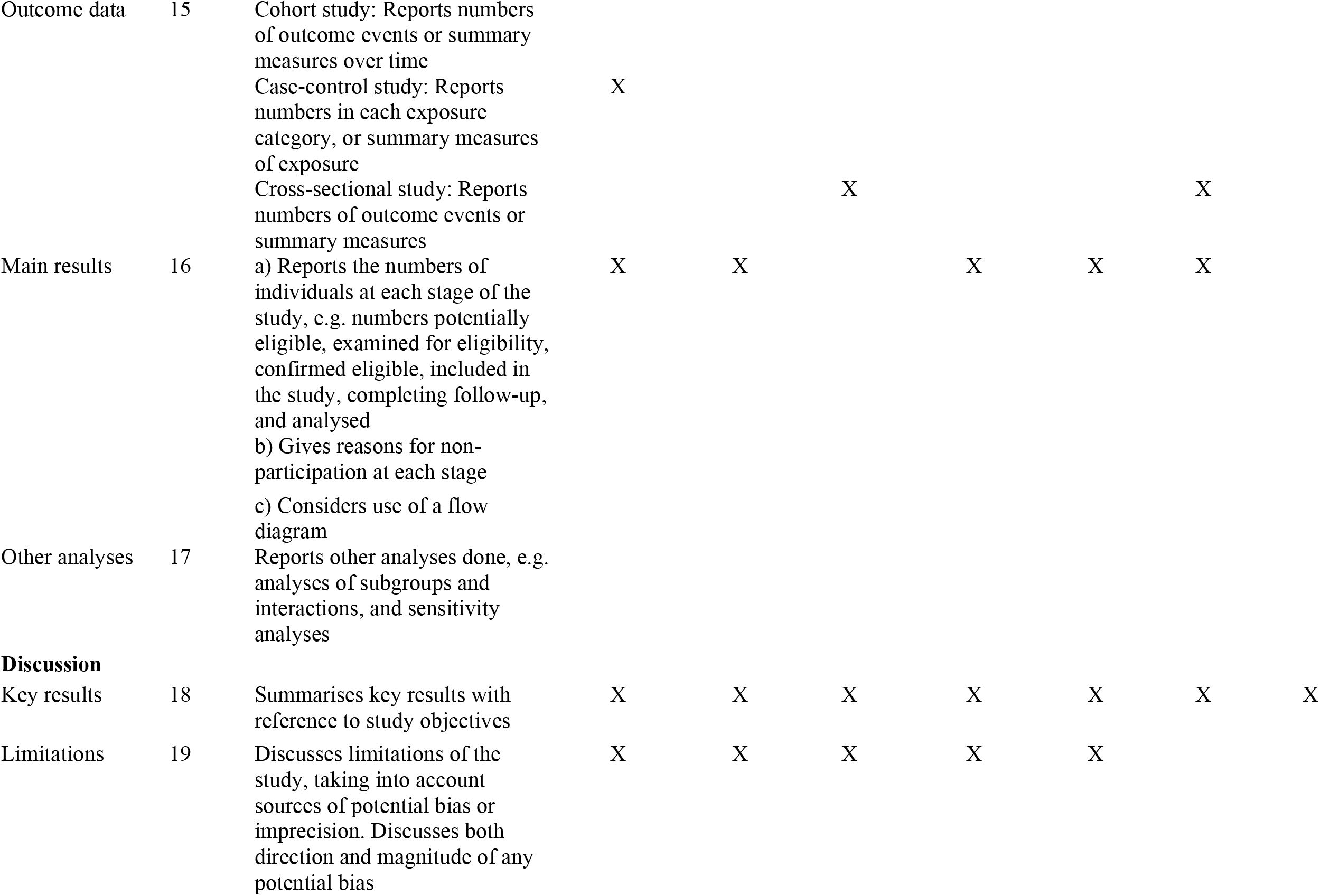

|  |  |  |  |  |  |  |  |  |  |
| --- | --- | --- | --- | --- | --- | --- | --- | --- | --- |
| Outcome data | 15 | Cohort study: Reports numbers of outcome events or summary measures over time<br>Case-control study: Reports numbers in each exposure category, or summary measures of exposure<br>Cross-sectional study: Reports numbers of outcome events or summary measures | X |  | X |  |  | X |  |
| Main results | 16 | a) Reports the numbers of individuals at each stage of the study, e.g. numbers potentially eligible, examined for eligibility, confirmed eligible, included in the study, completing follow-up, and analysed<br>b) Gives reasons for non-participation at each stage<br>c) Considers use of a flow diagram | X | X |  | X | X | X |  |
| Other analyses | 17 | Reports other analyses done, e.g. analyses of subgroups and interactions, and sensitivity analyses |  |  |  |  |  |  |  |
| **Discussion** |  |  |  |  |  |  |  |  |  |
| Key results | 18 | Summarises key results with reference to study objectives | X | X | X | X | X | X | X |
| Limitations | 19 | Discusses limitations of the study, taking into account sources of potential bias or imprecision. Discusses both direction and magnitude of any potential bias | X | X | X | X | X |  |  |

**Table I.**
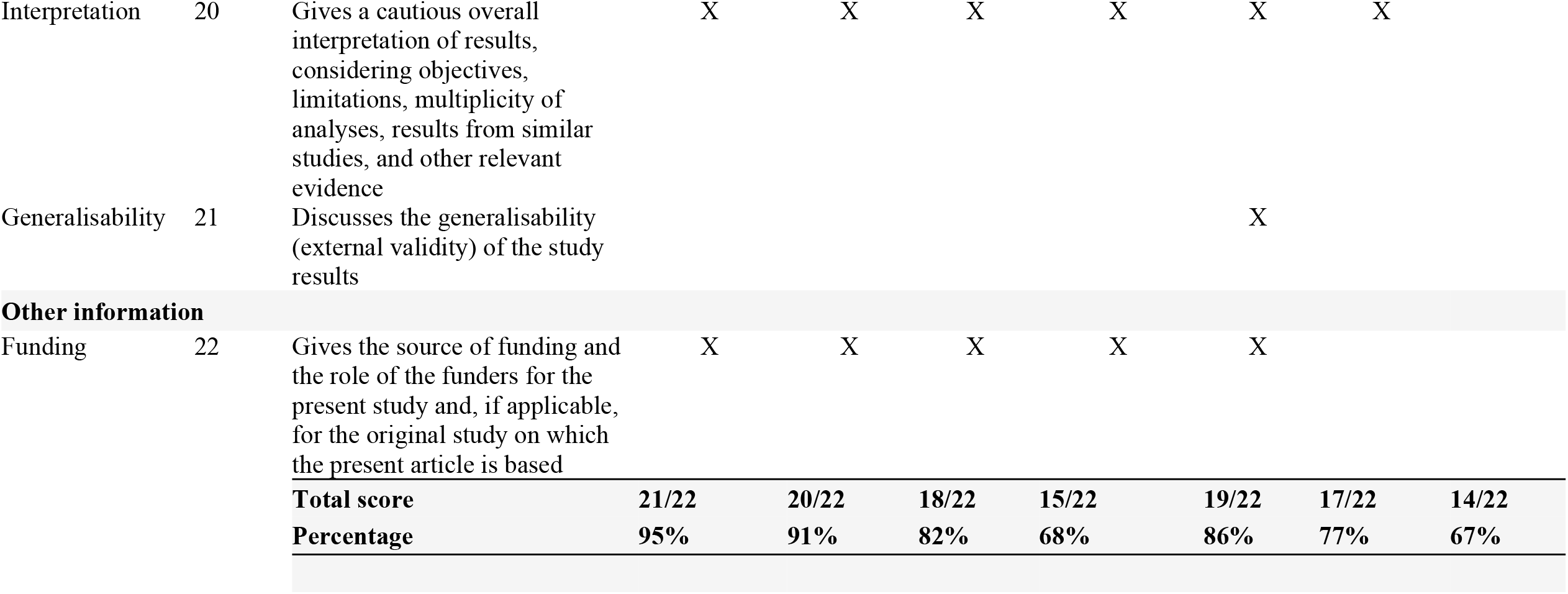

